# Endothelial Microvesicles: A Plasma Biomarker of Futile Recanalization Post Intravenous Thrombolysis in Patients with Stroke

**DOI:** 10.64898/2026.06.25.26356528

**Authors:** Huiting Zhang, Xiaobing Xu, Zhiwen Zhou, Yanyu Chen, Zhimin Liao, Jiayuan Wu, Junding Xian, Wangtao Zhong, Xiaotang Ma

## Abstract

Investigating futile recanalization indicators is very important. Here, we explored plasma endothelial microvesicles (EMVs) as biomarkers for recombinant tissue plasminogen activator (rtPA)-treated acute ischemic stroke. This study prospectively enrolled 195 acute IS patients who underwent rtPA and measured plasma EMVs levels via fluorescence nanoparticle tracking analysis at baseline, 24 h and 90 days. Early futile recanalization was assessed by transcranial Doppler and the National Institutes of Health Stroke Scale. The ROC curves and corresponding areas under the curve (AUC) of the EMVs were analysed. The plasma EMVs levels at baseline and 24 h were positively related to both early and late futile recanalization. In both the late recanalization and futile recanalization groups, the plasma levels of EMVs significantly increased at 24 h but decreased at 90 days. For early futile recanalization, the baseline and 24-h EMVs AUCs were 0.7 and 0.67, respectively. For late futile recanalization, the AUCs for baseline and 24-h EMVs levels were 0.52 and 0.66, respectively. Collectively, the results imply that the plasma level of EMVs could serve as a surrogate indicator of futile recanalization (both early and late) following rtPA administration in acute IS.

**Clinical Trial Registration:** https://www.chictr.org.cn; Number: ChiCTR2300074468

## 1. Introduction

Stroke is the foremost contributor to disease-attributed death globally [1]. Ischemic stroke (IS), which arises from obstruction of cerebral blood vessels, accounts for the majority of stroke cases [2]. Recanalization and restoration of blood flow are closely related to the clinical prognosis of IS patients [3]. The use of intravenous rtPA within a 4.5-hour time window is endorsed as an efficacious thrombolytic strategy for acute IS (AIS) [4]. Currently, transcranial Doppler, digital subtraction angiography and computed tomographic angiography are prominent methods for evaluating the recanalization status [5, 6]. Nevertheless, these examinations are either associated with potential risks, such as renal function impairment and ionizing radiation exposure, or are unable to fully reflect the true recanalization and prognosis of patients [7–9]. More than 50% of patients experience poor clinical outcomes despite the confirmation of complete vascular recanalization using the aforementioned methods; this condition is known as futile recanalization [10, 11]. Exploring biomarkers to evaluate thrombolytic efficacy might be more personalized and could provide guidance for subsequent endovascular thrombectomy.

Vascular homeostasis relies heavily on the endothelium, which resides at the interface of blood and interstitial tissues [12]. Impairment of endothelial cell (EC) constitutes an early pathophysiological feature of IS and contributes to poor recanalization and an unfavourable prognosis after intravenous thrombolysis [13]. Endothelial microvesicles (EMVs) are phospholipid microvesicles of 100–1000 nm released from ECs in response to activation and stress conditions [14]. EMVs are emerging as biomarkers for EC dysfunction-related diseases, including coronary heart disease, hypertension, and cerebrovascular diseases [15]. Plasma EMVs are detectable in the very early disease phase, for example, plasma EMVs levels increase significantly within 3 minutes in patients with intermediate coronary lesions [16]. Our group previously reported that EMVs could be used as indicators of EC damage and that the highest plasma levels of EMVs were detected in IS patients with large artery occlusion [14]. Fan and Huo et al. reported that a positive association existed between plasma EMVs concentrations and the incidence of acute coronary syndrome and poor cardiovascular outcomes [17]. Therefore, we suggest that EMVs might be biomarkers for the recanalization and prognosis of AIS after rtPA treatment.

In this study, the relationship between plasma EMVs and recanalization of AIS patients was analysed, and the plasma EMVs in AIS by subtype and severity were comparied. This study might introduce new perspectives on post-rtPA recanalization evaluation in AIS.

## 2. Methods

### 2.1 Study design and participants

This study was performed at the Affiliated Hospital of Guangdong Medical University (China). Patients were recruited between December 1, 2021, and December 1, 2023, eligible participants were included during this period and followed up for 90 days. The last enrolled patient completed the 90day follow-up on March 1, 2024. The study was in accordance with the STROBE guidelines and the ethical standards of the institutional/national research committee, as well as the Declaration of Helsinki (1964, including later amendments). The study protocol was approved by the hospital’s Ethics Committee (Approval number: PJ2021-092), and was registered in Chinese Clinical Trial Registry: https://www.chictr.org.cn; Number: ChiCTR2300074468.

Patients with AIS who received rtPA therapy were screened in this study. The following inclusion criteria were applied: 1) all participants were diagnosed with IS based on the ischemic stroke diagnostic criteria (2021) [18]; 2) Ischemic stroke causing measurable neurological impairment; 3) time from symptom onset to treatment <4.5 hours; 4) aged 18–80 years; 5) treatment with 0.9 mg/kg intravenous rtPA (alteplase; Boehringer Ingelheim, Ingelheim am Rhein, Germany) and no more than 90 mg, as recommended by the instructions; and 6) Written informed consent was secured from all participants and/or their legally authorized representatives.. The exclusion criteria were as follows: 1) mild or rapidly resolving stroke symptoms; 2) pregnancy; 3) Seizure at onset leaving residual neurological impairments; 4) major surgery or serious trauma in the preceding 14 days; 5) recent gastrointestinal or urinary tract haemorrhage (within 21 days); 6) acute myocardial infarction within the last 3 months; 7) serious liver and kidney diseases; and 8) severe stroke, defined as a National Institutes of Health Stroke Scale (NIHSS) score ≥ 25 (range 0-42, higher scores denote greater neurological severity). The timeline of the study was shown in Figure 1.

**Figure 1.**
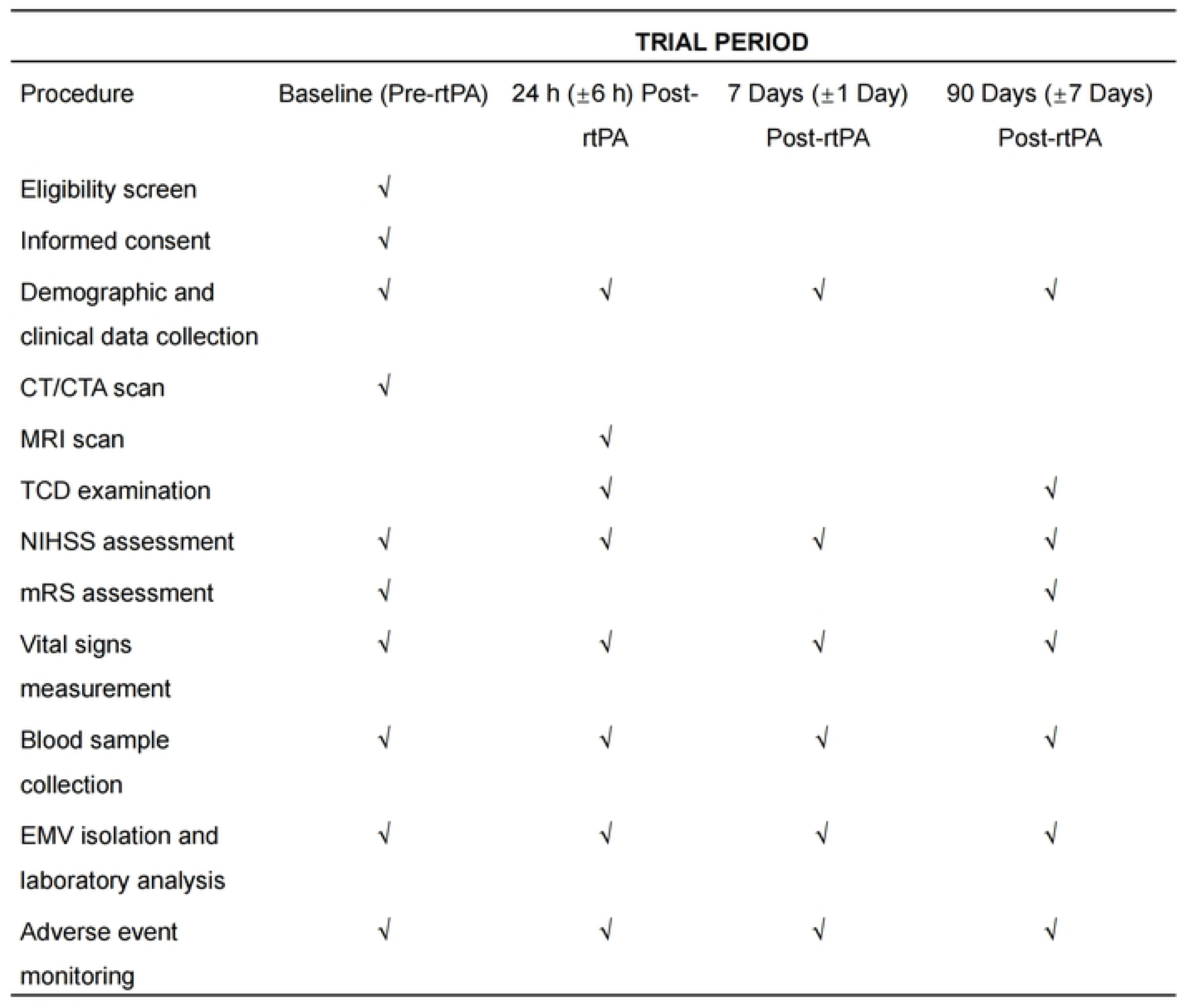
Study timeline.

### 2.2 Sample size calculation

The sample size was calculated based on the ROC curves and corresponding areas under the curve (AUC) of the EMVs in early futile recanalization. In our preliminary study, the AUC of EMVs for predicting early futile recanalization was 0.65, with the null hypothesis AUC set at 0.5. With power set at 80% and an α level at 0.05, the estimated sample size was 114 patients. Adding 20% loss to follow-up, the sample size estimate was increased to 143 patients. This estimate was conducted using PASS software (NCSS, LLC. Kaysville, Utah, USA) version 15.0. Standardized data collection protocols were employed, research personnel were trained, and follow-up communications were arranged to increase compliance.

### 2.3 Collection of clinical information

For each patient, we collected demographic and laboratory data (detailed in the Supplementary Materials), and related scales, including the National Institutes of Health Stroke Scale (NIHSS: 0-42, higher scores reflect more severe neurologic deficits, were assessed at admission and at 24 h, and the modified Rankin scale (mRS; global disability, 0-6) was measured at admission and at 90 days [19]. The Alberta Stroke Program Early Computed Tomography Scores (ASPECTS; 0-10, lower values indicating a larger infarct area) [20]. The scales were recorded by trained neurologists unknown to the other clinical data.

### 2.4 Outcome measurement

Primary end point was early futile recanalization. The secondary endpoints were late futile recanalization and the correlations between plasma EMVs levels and the severity, infarct volume, and subtypes of IS. Early futile recanalization was defined as complete recanalization confirmed by transcranial Doppler (TCD) with thrombolysis in brain ischaemia (TIBI) grades 4–5 and without a neurological improvement at 24 h (≥4 point reduction in the NIHSS score) after rtPA therapy [21]. Late futile recanalization was defined as complete recanalization tested by TCD (TIBI grade 4–5) with an mRS score ≥ 2 at 90 days following rtPA therapy [22].

### 2.5 Transcranial Doppler (TCD) assessment

Recanalization was assessed using TCD (Elica, EMS-9PB, China) at 24 h and 90 days after rt-PA therapy according to the standard operating procedure. Intracranial vessel residual flow signals was assessed by the thrombolysis in brain ischemia (TIBI) grading system..TIBI waveforms were graded from 0-5: 0, absent; 1, minimal; 2, blunted; 3, dampened; 4, stenotic; and 5, normal [23]. Complete recanalization was confirmed if the TIBI flow improved to grade 4 or 5 and TIBI flow grade 2 or 3 was defined as partial recanalization. Grade 0–1 TIBI cerebral blood flow was considered to indicate no recanalization [24].

### 2.6 IS severity and infarct volume measurements

The NIHSS score and infarct volume were used to evaluate stroke severity. NIHSS scores ≤ 5 and > 5 were defined as minor and moderate–severe stroke, respectively [25]. The infarct volume was determined by magnetic resonance imaging (MRI), as previously recommended [26]. Briefly, patients were scanned with a 3.0 T MRI (Philips Medical Systems). The total lesion volume was calculated as the highly hyperintense signal area per layer multiplied by the number of layers in accordance with the Pullicino formula (length × width × number of layers/2). IS patients were divided into small (<5 cm^3^) and moderate–large infarct groups (≥5 cm^3^), as previously recommended [14].

### 2.7 Identification and measurements of plasma EMVs

Plasma EMVs were isolated by density gradient centrifugation and measured by nanoparticle tracking analysis (NTA). The character of the EMVs were determined by performing transmission electron microscopy (TEM), as previously reported [14]. Western blotting was applied to the presence of Annexin V, a specific marker in EMVs. [27]. The technicians who assessed the EMVs level were masked to the clinical characteristics. and the recanalization status of the subjects. Detailed information can be found in the Supplemental Materials.

### 2.8 Statistical analysis

GraphPad Prism 7.0 and SPSS 22.0 were employed for all statistical calculations. Continuous data were expressed as mean ± standard deviation (SD) or median with interquartile range (IQR). Normality testing was carried out using the D’Agostino Pearson omnibus test. Comparisons of continuous variables were performed via one-way ANOVA, followed by either the Mann-Whitney U test or Student’s t-test, as appropriate. Categorical variables were analyzed with the chi-square test or Fisher’s exact test. Pearson’s correlation was used to evaluate the relationship between EMVs levels and both the NIHSS score and the 90-day mRS score. To explore the potential independent role of EMVs in predicting early and late futile recanalization, multivariate logistic regression with stepwise variable selection was applied. Results were reported as adjusted odds ratios (ORs) along with their 95% confidence intervals (CIs). Receiver operating characteristic (ROC) curves were generated to determine the area under the curve (AUC), thereby assessing the sensitivity and specificity of EMVs for diagnosing futile recanalization after rtPA therapy. The handling of missing data will follow the principles specified in the ICH-E9 and the CPMP/EWP/1776/99Rev1.1 Guideline on Missing Data in confirmatory trials Guidelines. A P-value of less than 0.05 was regarded as statistically significant.

## 3. Results

### 3.1 Characteristics of the subjects

From December 1st, 2021, to December 1st, 2023, a total of 204 acute IS patients were screened at the hospital. Among the excluded patients, two patients had severe renal insufficiency, two patients did not provide informed consent, and five patients refused blood collection. A cohort of 195 patients was included for assessment of plasma EMVs’ role in early futile recanalization. Eleven patients did not complete the 90-day followup. A flowchart of the present study is provided in Figure 2. Fifty-one (26.2%) patients achieved complete or partial recanalization, and 144 (73.8%) experienced early futile recanalization after rtPA treatment. Baseline demographic and clinical profiles of all enrolled patients are presented in Table 1. The median interval from stroke symptom onset to study inclusion was 3 hours (interquartile range: 2-4 hours) for both the early recanalization cohort and the futile recanalization cohort. The duration until the initiation of rtPA therapy was 38.8±12.2 minutes in the early recanalization group and 38.6±14.2 minutes in the early futile recanalization group. The median NIHSS and ASPECT scores were 4 (interquartile range, 2 to 8) and 9 (interquartile range, 8 to 10), respectively, in the early recanalization group and 6 (interquartile range, 2 to 13) and 9 (interquartile range, 8 to 9), respectively, in the early futile recanalization group. The proportions of anterior circulation infarcts in patients with early recanalization and futile recanalization were 82.4% and 79.9%, respectively. The level of LPA in the early futile recanalization group was significantly exceeded tthat in the recanalization group (*p*=0.03). Other baseline demographic and clinical parameters were well balanced between the two study groups (Table 1). As shown in Supplementary Table 1, the secondary end points at 90 days were obtained for 184 patients, and the baseline characteristics of the late recanalization group were collected. A total of 73 (36.7%) and 111 (60.3%) patients achieved late recanalization and late futile recanalization, respectively, after rtPA treatment. The characteristics of the late recanalization and futile recanalization groups did not differ significantly.

**Figure 2.**
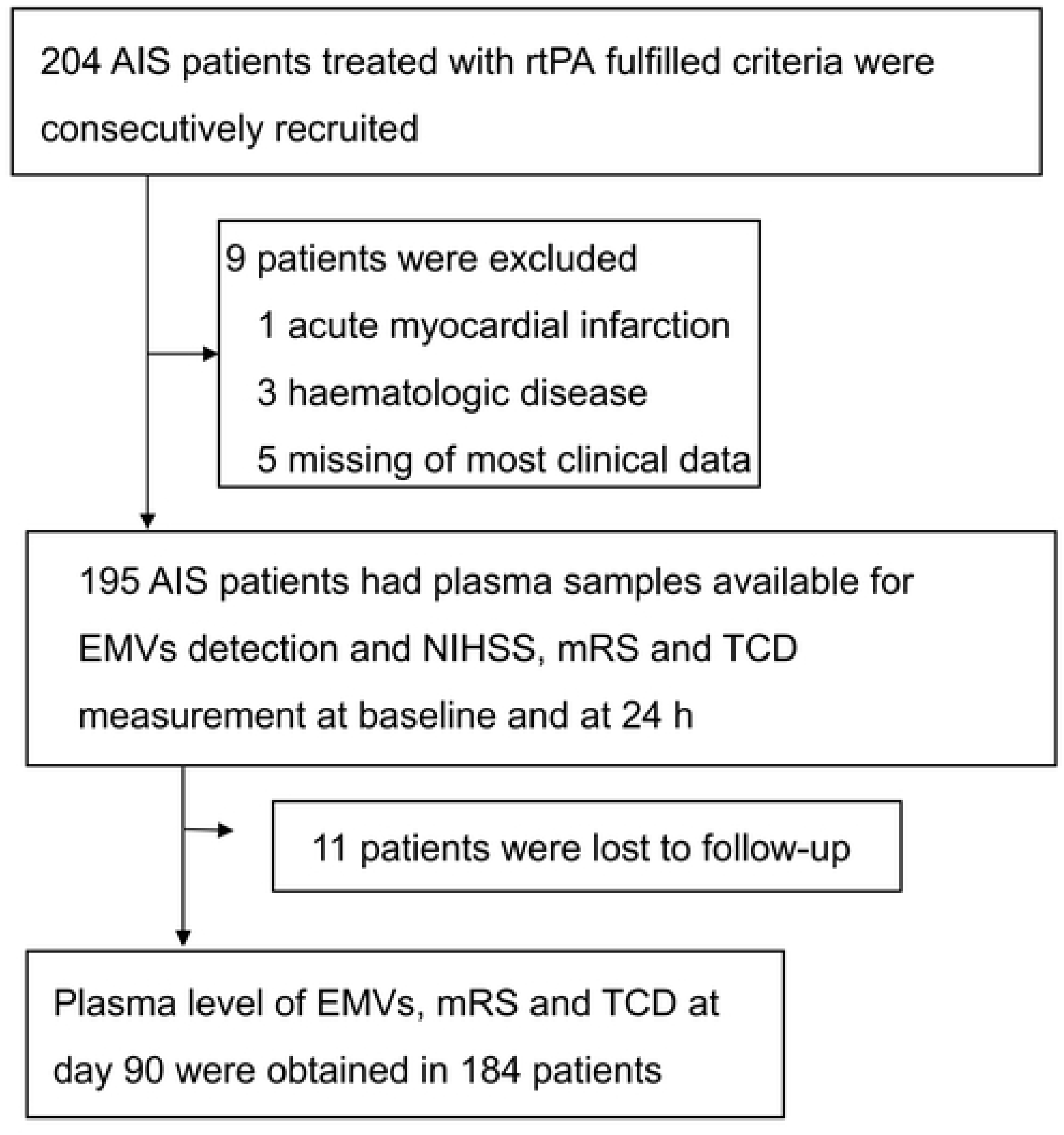
Flowchart of this study. Abbreviations: AIS, acute ischemic stroke; rtPA, recombinant tissue plasminogen activator; EMVs, endothelial microvesicles; NIHSS, National Institutes of Health Stroke Scale; mRS, modified Rankin scale; TCD, transcranial Doppler

### 3.2 Characterization of isolated EMVs

As illustrated in Supplementary Figure 1, TEM images and NTA showed that the sizes of the EMVs obtained from IS patients at baseline, 24 h, and 90 days displayed diameters from 100 nm up to 600 nm, and the sizes of these EMVs were not significantly different (Supplementary Figure 1A, B). Using western blot analysis, we confirmed the expression of a specific marker (Annexin V) in EMVs (Supplementary Figure 1C). The efficiency of EMVs (defined as CD105^+^CD144^+^ MVs) was tested by fluorescence NTA, which revealed that approximately 83% of the EMVs were isolated and purified and that the purification of the EMVs was not statistically significantly different (Supplementary Figure 1D, E).

### 3.3 Endpoint analysis

In terms of the primary endpoint, univariate analysis found that both baseline and 24-hour plasma EMV levels were significantly associated with early futile recanalization (baseline EMVs levels: OR=0.59, 95% CI: 0.42–0.83, *p*=0.002; 24-h EMVs levels: OR=0.78, 95% CI: 0.65–0.93, *p*=0.006; Table 2). Both baseline and 24 h plasma EMV levels (baseline EMVs levels: adjusted OR=0.41, adjusted 95% CI: 0.42–0.84, p=0.003; 24-h EMVs levels: adjusted OR=0.79, 95% CI: 0.66–0.94, *p*=0.008; Table 2). were independently correlated with early futile recanalization following adjustment for admission ASPECTS, baseline NIHSS score, age, systolic blood pressure (SBP), lipoprotein phospholipase A (LPA) level, homocysteine (HCY) level and cholesterol (CHOL) level. With respect to the secondary end points, univariate and multivariate analyses revealed that the plasma levels of EMVs at baseline and 24 h were significantly related to late futile recanalization (baseline EMVs levels: OR=0.74, 95% CI: 0.56–0.89, *p*=0.03; adjusted OR=0.72, adjusted 95% CI: 0.54–0.95, p=0.02; 24-h EMVs levels: OR=0.85, 95% CI: 0.74–0.99, *p*=0.04; adjusted OR=0.84, adjusted 95% CI: 0.70–0.99, p=0.04; Supplementary Table 2).

A positive correlation between baseline plasma EMV levels and the baseline NIHSS score was identified via correlational analysis (R=0.34, 95% CI: 0.21–0.46, *p*<0.001; Supplementary Table 3), 24-h NIHSS score (R=0.33, 95% CI: 0.2–0.45, *p*<0.001; Supplementary Table 3) and 90-day mRS score (R=0.28, 95% CI: 0.14–0.41, *p*<0.001; Supplementary Table 3). The EMVs levels at 24 h were positively correlated with the 24-h NIHSS score (R=0.41, 95% CI: 0.28–0.52, *p*<0.001; Supplementary Table 3) and 90-day mRS score (R=0.16, 95% CI: 0.02–0.3, *p*=0.03; Supplementary Table 3). The EMVs levels at 90 days were positively correlated with the 90-day mRS score (R=0.23, 95% CI: 0.09–0.36, *p=*0.002; Supplementary Table 3). As shown in Figure 3, the plasma levels of EMVs at baseline and 24 h were dramatically increased in the early futile recanalization compared with the recanalization groups (baseline EMVs levels: 3.67±1.36 *vs.* 2.79±1.05, *p*<0.001; 24-h EMVs levels: 5.47±2.59 *vs.* 4.21±1.78, *p*=0.001; Figure 3A). The difference in the plasma level of EMVs at baseline was not statistically significant between the late recanalization and late futile recanalization groups (3.43±1.28 *vs.* 3.44±1.50, *p*=0.97; Figure 3B). The EMVs levels at 24 h and 90 days were substantially increased in the late futile recanalization group compared with the recanalization group (24-h EMVs levels: 5.55±2.54 *vs.* 4.19±2.06, *p*=0.0002; 90-day EMVs levels: 3.41±1.88 *vs.* 1.86±0.97, *p*<0.0001; Figure 3B). The plasma levels of EMVs were highest at 24 h than at baseline and 90 days in the late futile recanalization group (24-h EMVs levels *vs.* baseline EMVs levels: 5.55±2.54 *vs*. 3.43±1.28, *p*<0.001; 24-h EMVs levels *vs.* 90-day EMVs levels: 5.55±2.54 *vs*. 3.41±1.88, *p*<0.001, Figure 3B). Results indicated a lack of statistically significant difference in plasma EMV concentrations between baseline and 90 days in the late futile recanalization. In the late recanalization group, the 90-day EMVs levels were dramatically decreased compared with the baseline EMVs levels and 24-h EMVs levels (baseline EMVs levels *vs.* 90-day EMVs levels: 3.44±1.50 *vs*. 1.86±0.97, *p*<0.001; 24-h EMVs levels *vs.* 90-day EMVs levels: 4.19±2.06 *vs*. 1.86±0.97, *p*<0.0001; Figure 3B).

**Figure 3.**
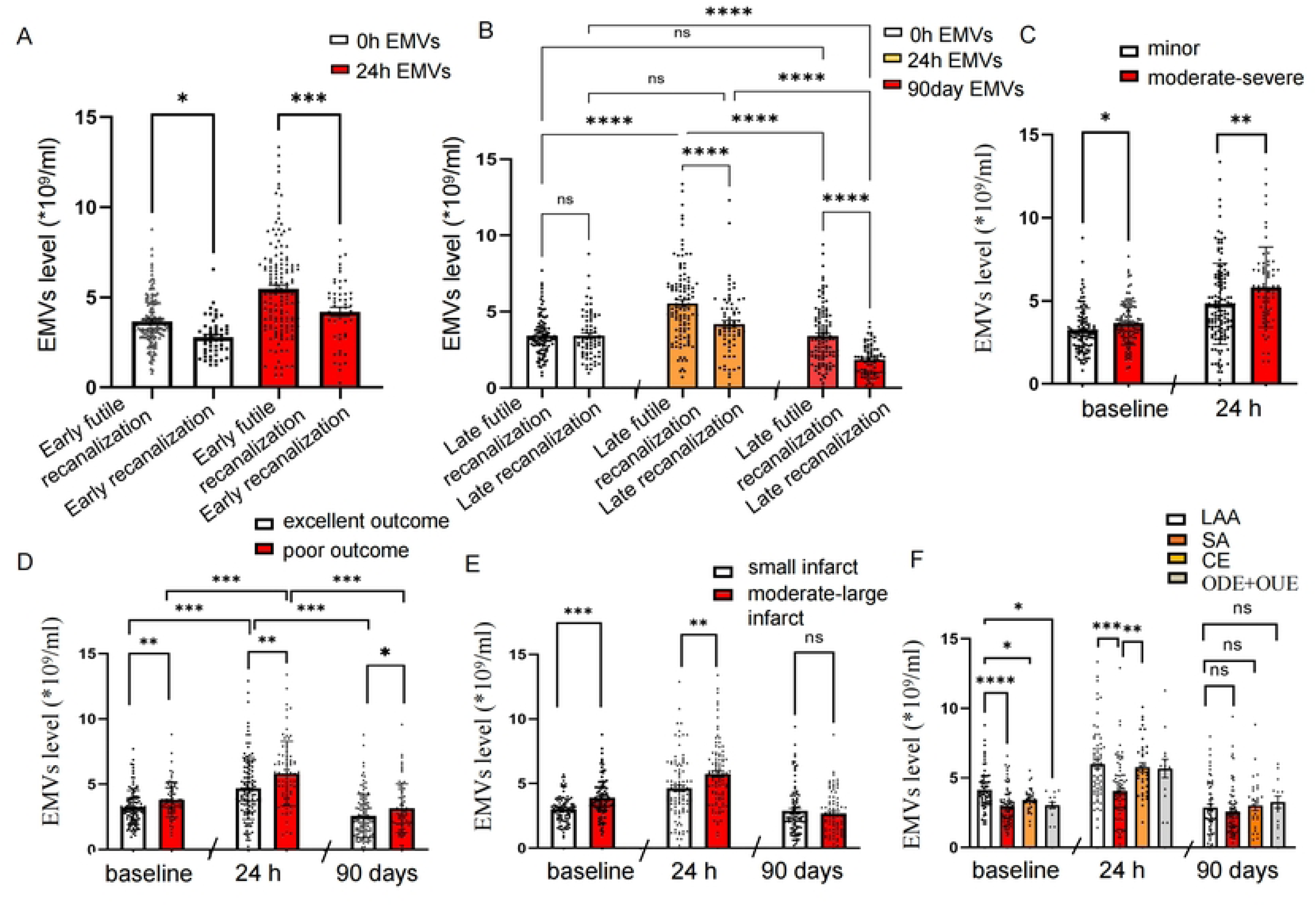
Analysis of plasma EMVs levels at different time points in patients with early and late futile recanalization and with different IS severities and TOAST classifications. (**A**) NTA results showing that the plasma levels of EMVs at baseline and 24 h were obviously increased by approximately 31% and 30%, respectively, in IS patients in the early futile recanalization group compared with those in the recanalization group (Mann–Whitney U test; \**p*=0.01 and \*\*\**p*<0.001). (**B**) The plasma levels of EMVs at 24 h and 90 days in late futile recanalization patients were approximately 1.3-fold and 1.8-fold higher than those in late recanalization patients (Mann–Whitney U test, \*\*\**p*<0.001), and the baseline EMP levels in late futile recanalization and late recanalization patients were not significantly different. The plasma EMVs level significantly increased at 24 h but decreased at 90 days in both the late futile recanalization group and the late recanalization group (Kruskal–Wallis ANOVA, \*\*\**p*<0.001). (**C**) Plasma levels of EMVs were clearly higher in the moderate–severe group than in the minor group at baseline and 24 h (baseline: Mann–Whitney U test, \**p*=0.02; 24 h: Mann–Whitney U test, \*\**p*=0.008). (**D**) The plasma levels of EMVs increased in the groups with poor outcomes at baseline, 24 h and 90 days (baseline: Mann–Whitney U test, \*\**p*=0.002; 24 h: Mann–Whitney U test, \*\**p*=0.001; 90 days: Mann–Whitney U test, \**p*=0.03). (**E**) The results revealed that the plasma levels of EMVs were clearly increased in the moderate–large infarct groups at baseline and 24 h (baseline: Mann–Whitney U test, \*\*\**p*<0.001; 24 h: Mann–Whitney U test, \*\**p*=0.002), and the comparison of 90-day EMVs levels between the small and moderate–large infarct groups showed no significant differences. (**F**) The plasma levels of EMVs in the LAA subgroup were higher than those in the other subgroups at baseline and 24 h. (baseline: Kruskal–Wallis ANOVA, LAA *vs*. SA, \*\*\**p*<0.001; LAA *vs*. CE, \**p*=0.03; LAA *vs*. ODE+OUE, \**p*=0.02; 24 h: Kruskal–Wallis ANOVA, LAA *vs*. SA, \*\*\**p*<0.001; CE *vs*. SA, \*\**p*=0.002). At 24 h, significant differences in the plasma EMVs levels were not observed among the LAA, CE and ODE+OUE subgroups. The plasma EMVs levels at 90 days were not significantly different among the LAA, SA, CE and ODE+OUE groups. The data are presented as the means±SDs.

IS severity was determined by measuring the NIHSS score and infarct volume. As presented in Figure 3C, IS patients treated with rtPA were divided into minor (NIHSS score≤5: baseline, n=99; 24 h, n=96) and moderate–severe (NIHSS score>5: baseline, n=133; 24 h, n=62) IS according to the NIHSS score. The plasma EMVs level at baseline in the moderate–severe group was approximately 1.14 times higher than that in the minor group (Figure 3C). The plasma level of EMVs clearly elevated by 20% in the moderate–severe group compared with that in the minor group at 24 h (Figure 3C). Moreover, the baseline, 24-h and 90-day EMVs levels were dramatically increased in the poor outcome groups (mRS score of 2–6) in comparism to the excellent outcome groups (mRS score of 0–1) (Figure 3D). As shown in Figure 3E, the infarct volume in patients with IS was measured using MRI. The numbers of small infarcts were 101/195 (at baseline and 24 h) and 100/184 (at 90 days), and the numbers of moderate–large infarcts were 94/195 (at baseline and 24 h) and 84/184 (at 90 days). The results revealed that the plasma EMVs levels at baseline and 24 h were dramatically higher in the moderate–large infarct groups than in the small infarct groups (Figure 3E). The 90-day EMVs levels in the small and moderate–large infarct groups were not significantly different (Figure 3E). As shown in Figure 3F, 65 patients had LAA, 79 had SA, 37 had CE and 14 had ODE+OUE at baseline and 24 h, and 63 patients had LAA, 79 had SA, 28 had CE and 14 had ODE+OUE at 90 days according to the TOAST classification. The plasma EMVs level at baseline in the LAA group was significantly higher than those in the SA, CE, and ODE+OUE groups (Figure 3F). The comparisons among the SA, CE and ODE+OUE groups did not show statistically significant differences. At 24 h, the levels of EMVs were higher in the LAA and CE groups than in the SA group (Figure 3F). No statistical differences were shown among the LAA, SA, CE and ODE+OUE groups at 90 days.

### 3.4 Plasma EMVs levels showed significant diagnostic value for early and late futile recanalization after rtPA therapy

ROC curves were constructed to discriminate IS patients who experienced early and late futile recanalization from IS patients who experienced recanalization to evaluate the diagnostic value of plasma EMVs levels for early and late futile recanalization after rtPA treatment. As shown in Figure 4, Table 3, and Supplementary Table 4, the areas under the curve (AUCs) for baseline and 24-h EMVs levels to determine early futile recanalization were 0.7 and 0.67, respectively, and both were higher than the AUC of the NIHSS score (0.58). The diagnostic thresholds of the baseline and 24-h EMVs levels were 3.25×10^9^/ml (sensitivity 58.3%, specificity 74.5%, 95% CI: 0.62–0.78, *p*<0.001; Figure 4A and Table 3) and 5.98×10^9^/ml (sensitivity 40.4%, specificity 88.2%, 95% CI: 0.59–0.75, *p*=0.004; Figure 4A and Table 3), respectively. The sensitivity and specificity of the NIHSS score for diagnosing early futile recanalization were 54.2% and 64.7%, respectively. The joint predictor of baseline EMVs levels, 24-h EMVs levels and the NIHSS score obtained higher diagnostic efficacy. As illustrated in Figure 4B and Supplementary Table 4, the AUCs for baseline EMVs levels and 24-h EMVs levels to assess late futile recanalization were 0.52 and 0.66, respectively, and both were higher than the AUC of the NIHSS score (0.5). The diagnostic thresholds of the baseline EMVs levels and 24-h EMVs levels were 3.5×10^9^/ml (sensitivity of 42.3% and specificity of 65.8%, 95% CI: 0.43–0.61, *p*=0.04) and 5.87×10^9^/ml (sensitivity of 44.7% and specificity of 86.3%, 95% CI: 0.59–0.71, *p*<0.0001), respectively. The sensitivity and specificity of the NIHSS score for the diagnosis of late futile recanalization were 49.5% and 58.9%, respectively. The joint predictor of baseline EMVs levels, 24-h EMVs levels, and the NIHSS score was the best, with an AUC of 0.69. These findings suggest that plasma EMVs levels might serve as reliable predictors for early and late futile recanalization after rtPA treatment and that combining EMVs levels and the NIHSS score could increase the diagnostic efficacy.

**Figure 4.**
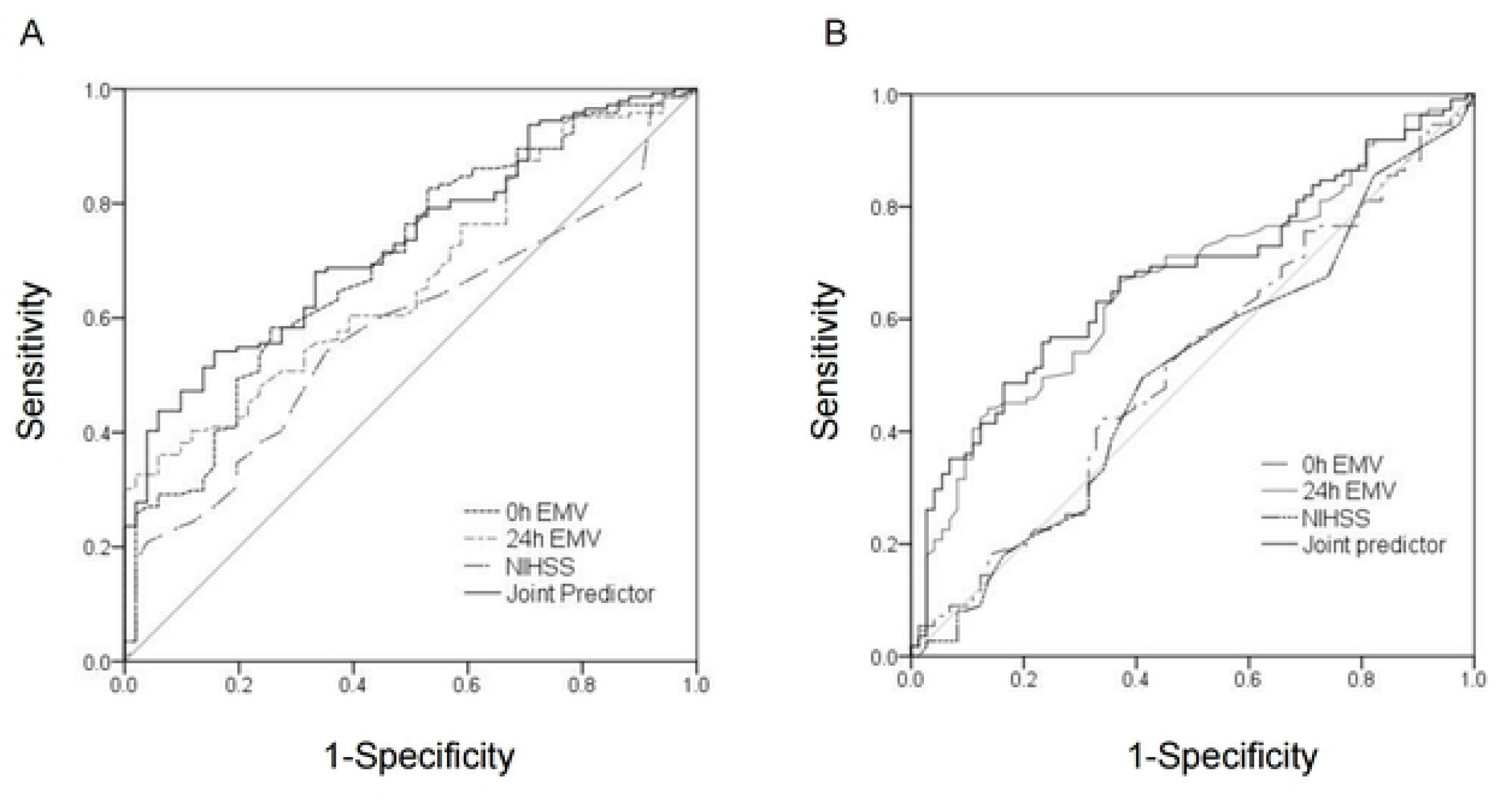
ROC curves for discriminating early and late futile recanalization using EMVs levels and the NIHSS score. (**A**) ROC curve for discriminating early futile recanalization based on the EMVs levels and the NIHSS score. The AUCs of both the baseline (0.7) and 24-h EMVs levels (0.67) were higher than that of the NIHSS score (0.58), and the AUC of their combination was 0.71. (**B**) ROC curve for discriminating late futile recanalization based on the EMVs levels and the NIHSS score. The AUCs of both the baseline (0.52) and 24-h EMVs levels (0.66) were higher than that of the NIHSS score (0.5), and the AUC of their combination was 0.69.

## 4. Discussion

Our findings revealed that IS patients with early and late futile recanalization following intravenous rtPA treatment exhibited a significant increase in circulating plasma EMVs. EMVs levels were associated with large artery atherosclerotic IS defined by the TOAST classification and were positively related to the NIHSS score and infarct volume. EMVs were identified as independent contributors to early and late futile recanalization after rtPA therapy and possessed diagnostic value, as evidenced by ROC curve analysis. These results imply that the plasma EMVs level is a promising indicator for early and late futile recanalization after rtPA treatment and for the severity of IS and might funciton as a reliable indicator for direct reperfusion therapy.

The great value of recanalization in occluded vessels has been demonstrated in patients with acute IS [28]. Digital subtraction angiography, magnetic resonance angiography, computed tomographic angiography and transcranial Doppler have been widely applied to evaluate the recanalization of blood vessels [29, 30]. Unfortunately, despite the patency of the blood vessels detected by these examinations, many IS patients still experience a poor prognosis, which is considered futile recanalization [31, 32]. Blood-based biomarkers might be more individualized and objective for evaluating the recanalization of IS patients [19]. Currently, little is known about revascularization markers in rtPA-treated patients with AIS. Increased plasma levels of a disintegrin-like and metalloproteinase with thrombospondin type 1 motif, member 13 (ADAMTS13) were reported to be associated with rtPA-induced recanalization. ADAMTS13 displayed a sensitivity of 68.8% and a specificity of 55.3% in predicting 2-hour recanalization. No significant association was observed between ADAMTS13 activity and long-term functional outcomes. Due to its relatively low specificity and failure to predict the long-term prognosis of patients after thrombolysis, the use of ADAMTS13 prompted us to explore novel plasma markers [33]. EC damage can cause poor collateral circulation, subacute reocclusion, microvascular injury, and compromised brain autoregulation, contributing to futile recanalization [34, 35]. Therefore, exploring a personalized biomarker associated with EC function to determine the recanalization status may provide valuable information for physicians and patients to make optimized decisions, such as when they receive arterial thrombolysis or endovascular thrombectomy therapy after rtPA treatment.

EMVs are accepted as potential biomarkers of EC dysfunction and various diseases [17]. Previous studies have demonstrated that plasma levels of EMVs are augmented in patients with type 2 diabetes mellitus and hypertension [36]. Notably higher plasma EMV levels have been documented in acute myocardial infarction patients, which possess predictive value for major adverse cardiovascular events [37]. Our study is the first to identify a role for EMVs in predicting futile recanalization following thrombolysis and provides robust evidence to support the implementation of early targeted interventions for AIS. We found that the plasma levels of EMVs at baseline and 24 h were positively correlated with early futile recanalization, with the increase in 24-h EMVs levels being more pronounced. Moreover, baseline and 24-h EMVs levels exhibited predictive value for short-term futile recanalization following thrombolysis, with AUCs of 0.7 and 0.67, respectively. Plasma EMVs levels at 24 h and 90 days were positively correlated with late futile recanalization, and the increase in 24-h EMVs levels was more significant. The AUCs indicate the reliable diagnostic value of 24-h EMVs levels for late futile recanalization. These findings indicate that the plasma EMVs level at baseline could more effectively predict short-term futile recanalization after thrombolysis, whereas 24-h EMVs levels showed greater potential as a marker for predicting late futile recanalization, thereby enabling early intervention. Notably, the incorporation of 0-h EMVs levels and 24-h EMVs levels into a combined model with the NIHSS score significantly increased the predictive efficacy. Previous investigations have established the roles of individual factors such as baseline NIHSS score and ASPECTS in predicting reperfusion outcomes [38]. The clinical utility of this combined model is noteworthy for AIS management. In resource-constrained settings lacking access to sophisticated neuroimaging modalities (e.g., perfusion CT or MRI) are not readily available, the combined EMVs–NIHSS model offers a low-cost, rapid, and accessible alternative to guide clinical decision-making. For instance, patients with high baseline or 24-h EMVs levels and elevated NIHSS scores could be prioritized for urgent vascular intervention or intensive postthrombolysis monitoring, whereas those with favourable biomarker and clinical profiles might be candidates for less aggressive strategies, thereby reducing unnecessary healthcare costs.

In terms of functional change, the poor prognosis of IS patients treated with intravenous thrombolysis mostly manifests as inconsistent cerebral autoregulation (CA) and cerebral blood flow (CBF) [34]. Decreased CBF and collateral circulation are responsible for futile recanalization. EC dysfunction contributes to blood–brain barrier (BBB) breakdown, CBF dysregulation and CA insult and might further induce reperfusion injury, such as haemorrhagic transformation or bleeding [39, 40]. These previous reports suggest that EC dysfunction plays crucial roles in thrombosis and futile recanalization. Elevated EMVs levels can reflect EC damage, such as overwhelming apoptosis and oxidative stress [14]. The plasma EMVs possess positive correlation with the level of Von Willebrand factor, an important marker of EC dysfunction and a vital indicator of decreased CBF [41]. Therefore, we propose that the high EMVs level detected in patients with rtPA-induced futile recanalization is related to EC injury. In this study, we found that 90-day EMVs levels in the recanalization group decreased below the baseline level, whereas no statistical difference was shown in the futile recanalization group. These findings indicate that EC injury might have improved compared with the time of onset, contributing to successful recanalization. Consistent with our previous report [14], our results also revealed the associations of plasma EMVs levels with the acute phase, subtypes and severity of IS. The levels of oxidative stress markers such as homocysteine and malondialdehyde are substantially increased in IS patients [42, 43]. Increased levels of inflammatory factors such as interleukin (IL)-6, IL-1β, and tumour necrosis factor (TNF-α) and apoptosis were detected in IS patients and in middle cerebral artery occlusion (MCAO) rats [44]. Prestimulation with TNF-α in the human umbilical artery induced the production of EMVs, and EMVs could increase the reactive oxygen species (ROS) level [45]. These previous studies indicated that severe oxidative stress and apoptosis occur in patients with acute IS and that EMVs might be related to oxidative stress and are intimately involved in the process of IS. Furthermore, our data showed that the plasma EMVs levels were obviously increased in the moderate–severe and moderate–large infarct groups at baseline, 24 h and 90 days. The plasma levels of EMVs were positively correlated with the NIHSS and mRS scores. Severe and large infarct IS are also linked to oxidative stress and apoptosis. Relative to the mild group, patients with severe IS had significantly higher circulating ROS levels [14]. Reduced caspase-3 expression and increased bcl-2 expression were associated with better functional outcomes and smaller infarct volumes in MCAO mice [46]. Additionally, we observed the highest plasma EMVs levels in the LAA group at baseline and 24 h according to the TOAST classification. Oxidized low-density lipoprotein, which is a crucial oxidative indicator, was significantly correlated with death and poor clinical outcomes in the LAA subgroup of patients with IS [47]. EC apoptosis represents a critical initiating event in the development of atherosclerotic lesions [48]. Increased EC apoptosis has been observed in the endothelium of human atherosclerotic plaques [49]. These studies suggest that severe apoptosis and oxidative stress in the severe IS and LAA subgroups might induce high concentration of EMVs.

## 5. Conclusions

In conclusion, this novel study elucidates the role of plasma EMVs in thrombolytic efficacy. Patients with a high EMVs level might have a poor response to rtPA treatment and have a lower chance of achieving recanalization, and bridging therapy or direct mechanical thrombectomy might be further considered. This work not only expands the predictive tools for AIS but also provides a rationale for integrating molecular biomarkers into routine clinical assessments, paving the way for more precise and individualized stroke management. Several limitations of the study should be acknowledged. The sample size was relatively modest, and future multicentre, prospective studies with larger patient populations are warranted to validate the performance of the model. Generalizability of our findings may be limited by the singl center design in China, which may not reflect broader populations. The underlying mechanisms linking EMVs dynamics to recanalization outcomes require further exploration, particularly regarding the crosstalk between EMVs, BBB integrity, and postreperfusion inflammation.

## Data Availability

No data was generated by this study.

## Ethics statement

This study adhered to the Strengthening the Reporting of Observational Studies in Epidemiology (STROBE) guidelines and the ethical standards of the institutional and/or national research committee, complying with the 1964 Declaration of Helsinki and its later amendments. It was approved by the Ethics Committee of the Affiliated Hospital of Guangdong Medical University (Approval No. PJ2021-092) and registered with the Chinese Clinical Trial Registry (www.chictr.org.cn/), and the trial registration number is ChiCTR2300074468.

## Acknowledgements

None.

## Author Contributions

Huiting Zhang: Writing—review and editing, Writing—original draft, Visualization, Validation, Methodology, Investigation, Formal analysis, Data curation, Conceptualization. Xiaobing Xu: Writing—original draft, Methodology, Investigation, Formal analysis, Data curation. Zhiwen Zhou: Methodology, Investigation, Formal analysis, Data curation. Yanyu Chen: Data curation. Zhimin Liao: Investigation. Jiayuan Wu: Formal analysis. Junding Xian: Formal analysis. Wangtao Zhong: Writing—review and editing, Visualization, Validation, Investigation. Xiaotang Ma: Writing—review and editing, Visualization, Validation, Formal analysis, Data curation.

## Data availability statement

The data are provided within the manuscript or Supplementary Information files.

---

**Supplementary Figure 1.** Identification of EMVs from IS patients. (**A**) Representative images of EMVs detected using TEM. Scale bars: 100 nm. (**B**) Comparison of the sizes of the EMVs in plasma detected using NTA. The results showed that the sizes of the EMVs did not differ significantly (n=3, Kruskal–Wallis ANOVA, *p*=0.67). (**C**) The levels of an EMV-specific marker (annexin V) were detected by western blotting. (**D**) Representative plots showing the size/concentration distribution of plasma EMVs isolated from IS patients. CD105^+^ beads were used to isolate microvesicles (MVs) in fluorescence/nonfluorescent mode. Red curve: CD105^+^ MVs were measured in light scatter (nonfluorescence) mode. Orange curve: CD105^+^ MVs and CD144^+^ Q-dot–MVs were measured in fluorescence mode. (**E**) Efficiency of EMVs purification. The data showed no significant difference in the efficiency of the methods for purifying EMVs (n=3; Kruskal–Wallis ANOVA, *p*=0.077). EMVs, endothelial microvesicles; IS, ischemic stroke; TEM, transmission electron microscopy; NTA, nanoparticle tracking analysis.

